# Associations between participation in community arts groups and aspects of wellbeing in older adults in the United States: A propensity score matching analysis

**DOI:** 10.1101/2021.06.01.21258135

**Authors:** Jessica K. Bone, Daisy Fancourt, Meg E. Fluharty, Elise Paul, Jill K. Sonke, Feifei Bu

## Abstract

There is a social gradient in both arts engagement and wellbeing which may have led to an overestimation of the impact of arts engagement on wellbeing in previous research. Using data from 12,111 older adults in the Health and Retirement Study (2014-2016), we tested whether participation in community arts groups was associated with concurrent wellbeing. We measured life satisfaction (evaluative wellbeing), positive and negative affect (experienced wellbeing), and purpose in life, constraints on personal control, and mastery (eudaimonic wellbeing). We used propensity score matching to remove confounding by a range of demographic, socioeconomic, and health-related factors. Participating in arts groups was associated with higher positive affect (average treatment effect on the treated (ATT)=0.19, 95% CI=0.13-0.24), life satisfaction (ATT=0.10, 95% CI=0.05-0.16), purpose in life (ATT=0.08, 95% CI=0.02-0.14), and mastery (ATT=0.08, 95% CI=0.02-0.13) than not participating. Arts group participation was not associated with negative affect or constraints on personal control. After matching on a range of potential confounders, participation in arts groups was associated with the positive elements of evaluative, experienced, and eudaimonic wellbeing. Facilitating participation in community arts groups could help to promote healthy aging, enabling a growing segment of the population to lead more fulfilling and satisfying lives.

## Introduction

In recent years, definitions of healthy aging have been broadened beyond the simple absence of physical and mental health problems to emphasize the importance of wellbeing in several domains (1,2). Wellbeing is often divided into hedonic wellbeing, which relates to attaining pleasure and avoiding pain, and eudaimonic wellbeing, which relates to finding meaning and flourishing (3). Hedonic wellbeing can be further divided into evaluative wellbeing (life satisfaction) and experienced wellbeing (positive and negative affect; 4). In contrast, types of eudaimonic wellbeing include control, mastery, autonomy, and personal growth (5). The multidimensional structure of wellbeing reflects the range of priorities for healthy aging (6). Individuals who are happy and satisfied in later life may feel more autonomous and pursue more opportunities for development. Equally, these aspects of eudaimonic wellbeing may make people happier and more satisfied (6). There is also evidence that wellbeing is associated with better mental and physical health outcomes and higher social engagement in older adults, as well as longer life expectancies (7–12). Identifying ways to support wellbeing in older adults has thus been labelled a public health priority (13).

Arts, cultural, and creative activities provide a potential source of wellbeing as they can offer opportunities for cognitive stimulation, physical activity, social interactions, emotional bonding, collaborative learning, pursuit of collective goals, and developing self-esteem (14–17). There is strong evidence that engagement in the arts can enhance both hedonic and eudaimonic wellbeing (18). Systematic reviews of arts-based interventions (e.g. music, singing) have found reliable evidence for improvements in both types of wellbeing (18,19). More broadly, analyses of longitudinal data have demonstrated that cultural engagement (e.g., attending the theater, concerts, and museums), active arts participation (e.g., dance, music making, painting), and membership of community arts groups (e.g., education, arts, or music classes) are associated with greater subsequent evaluative, experienced, and eudaimonic wellbeing, over periods of up to ten years (20–25). In one study, cultural access was the second most important determinant of wellbeing in older adults, exceeding factors such as income, age, employment, and education (26).

However, previous research has a number of limitations. There is evidence for a social gradient in arts engagement, with factors such as income, education, and race/ethnicity associated with lower frequency of engagement and structural barriers to engaging in the arts (27,28). Previous studies have generally adjusted for these sociodemographic factors in ordinary least squares regression models. Not doing so could have led to an overestimation of the impact of arts engagement on wellbeing, as wellbeing may be similarly socially patterned (10,29). However, even after adjusting for potential confounders, residual imbalance between those who do and do not engage in the arts can still bias results (30). Some studies have employed more sophisticated methods to address this, such as fixed effects regression and propensity score matching, but have not found consistent evidence for the association between arts engagement and wellbeing in younger adults (23,31,32). There are very few studies of these associations specifically in older adults (20–22). Additionally, many studies have relied on a single-item measure of wellbeing which could lead to measurement issues in comparison to using multi-item questionnaires (23,25,31,33). Finally, most research has been based in the United Kingdom (UK) and Europe, limiting its generalizability to the very different social, cultural, and demographic contexts of the United States (US).

This study aimed to explore associations between participation in a local community arts group and concurrent evaluative, experienced, and eudaimonic wellbeing in older adults in the US. We used data from a large nationally representative cohort study of older adults in the US (the Health and Retirement Study) (34). To address the issue of confounding by demographic, socioeconomic, and health-related factors, data were analyzed using propensity score matching (PSM), which simulates a randomized trial with the measured covariates randomized between groups. To explore the concept of wellbeing in depth, we examined each domain of wellbeing using validated measures of life satisfaction (evaluative wellbeing), positive and negative affect (experienced wellbeing), and purpose in life, perceived constraints on personal control, and perceived mastery (eudaimonic wellbeing). We hypothesized that arts group participation would be associated with enhanced wellbeing across all domains.

## Methods

### Sample

Participants were drawn from the Health and Retirement Study (HRS); a nationally representative study of more than 37,000 individuals over the age of 50 in the US (34). The initial HRS cohort was interviewed for the first time in 1992 and followed-up every two years, with other studies and younger cohorts merged with the initial sample. Further details on study design are reported elsewhere (34). In this study, we combined five HRS public datasets (RAND HRS: Longitudinal File 2018 (V1); Detailed Imputations File 2018 (V1); 2014-2018 Fat Files).

We used data from HRS waves at which participation in a local community arts group was measured (2014-2016). Within each wave, 50% of participants were invited to an enhanced interview, meaning participants were eligible in either 2014 or 2016. Following this interview, the Psychosocial and Lifestyle Questionnaire was left behind for participants to complete and return by mail, which included questions on participation in community arts groups and wellbeing. In 2014, 9,549 were eligible and 7,541 (79%) participated. In 2016, 10,238 were eligible and 6370 (62%) participated. We restricted the sample to participants with complete data on participation in community arts groups, wellbeing outcomes, and all covariates. This produced a final sample size of 12,111 participants, 6,602 of whom participated in 2014 and 5,509 in 2016 (no participants completed both years).

All participants gave informed consent and this study has Institutional Review Board approval from the University of Florida (IRB201901792) and ethical approval from University College London Research Ethics Committee (project 18839/001).

### Exposure

Participants were asked how often they participated in a local community arts group such as a choir, dance, photography, theatre, or music group in the last month. Responses were recorded on a seven-point frequency scale, ranging from never to daily. We collapsed these responses into two categories, representing no participation (never/not in the last month) or participation at least once in the last month (once a month to daily).

### Outcomes

We analyzed six aspects of wellbeing, all of which were measured in the Psychosocial and Lifestyle Questionnaire. For evaluative wellbeing, life satisfaction was measured with the five-item Satisfaction with Life Scale (35,36). Scores ranged from one to seven, with higher scores indicating greater life satisfaction. For experienced wellbeing, positive affect was measured with a list of 13 single-word items describing affect during the last 30 days, and negative affect was measured with 12 single-word items. This measure was developed for HRS (37) and mostly included words from the Positive and Negative Affect Schedule Expanded Form (38). Scores ranged from one to five and higher scores indicated more positive or negative affect. For eudaimonic wellbeing, three scales were used. Purpose in life was measured using the seven-item purpose subscale of the Ryff Measures of Psychological Wellbeing (39). Scores range from one to six and higher scores indicated greater purpose in life. Two five-item measures of perceived control were included, the perceived constraints on personal control and perceived mastery scales (29). For both measures, scores ranged from one to six. For constraints, higher scores indicated more constraints (lower perceived control). For mastery, higher scores indicated greater mastery (higher perceived control).

We followed the HRS instructions on coding and index creation; for each outcome, summary scores were calculated as the average of responses to each item and were set as missing if responses on more than half of the items were missing (37). All outcomes were then standardized within our analytical sample (mean=0, standard deviation=1). A standardized score represents the number of standard deviations each participant’s raw score is from the overall mean of that measure.

### Covariates

We included demographic, socioeconomic, and health-related covariates. Based on their availability in the survey data, demographic covariates were age (years), gender (men, women), marital status (married, divorced/separated, widowed, never married), and race/ethnicity (White [including Caucasian], Black [including African American], Other [including American Indian, Alaska Native, Asian or Pacific Islander, Other]). In the public HRS data, more detailed information on race/ethnicity was collapsed to protect participant confidentiality, and this variable indicated the race/ethnicity as which participants primarily identified. Socioeconomic covariates were educational attainment (none, high school, college, postgraduate), employment status (employed, unemployed, disabled, retired, homemaker), total household income (in US Dollars), neighborhood safety (excellent, very good, good, fair, poor), and frequency of socializing with friends or family (<1 time a year, 1-2 times a year, every few months, 1-2 times a month, 1-2 times a week, ≥3 times a week).

Health-related covariates included difficulties relating to activities of daily living (ADLs). Participants were asked whether, because of a health or memory problem, they had any difficulty with dressing; bathing or showering; eating; getting in or out of bed; and walking across a room. A summary score was created to indicate the number of these activities with which participants had difficulty (40). We also measured number of difficulties relating to instrumental activities of daily living (IADLs): making phone calls; managing money; taking medications; shopping for groceries; and preparing hot meals. A variable describing long-term physical health conditions (none, one or more) indicated whether participants reported having diabetes, lung disease, cancer, heart conditions, high blood pressure, arthritis, complications from stroke, or other medical conditions. Finally, we included a measure of cognition, which was a summary of immediate and delayed recall test scores (range 0-20) (40).

### Statistical analysis

We investigated whether participating in community arts groups was associated with higher concurrent wellbeing. We aimed to address the issue that certain types of individuals may be more likely to participate in community arts groups by using propensity score matching (PSM). This involves estimating a propensity score for each participant, indicating how likely they are to participate in arts groups based on covariates. Propensity scores are then used to match individuals who participated in arts groups (the ‘treatment’ group) with those who did not participate (the ‘control’ group) (41,42). Matched participants should have almost identical distributions on all observed covariates, removing any confounding by these covariates. In this way, PSM simulates a randomized trial with the measured covariates randomized between groups. We used PSM to estimate the difference between the average outcome for arts group participants and the average outcome for the same group under the hypothetical scenario that they did not participate in arts groups (the average treatment effect on the treated; ATT).

Propensity scores were estimated using demographic, socioeconomic, and health-related covariates. Quadratic forms of continuous covariates were tested but there was no evidence (p>0.05) that these should be included. Where there was evidence that interactions between covariates improved the prediction of treatment (p<0.05), interaction terms were added (gender*marital status, gender*ADLs, race/ethnicity*marital status, marital status*long-term conditions, age*employment status, age*household income, age*cognition, neighborhood safety*long-term conditions, ADLs*IADLs). We assessed the propensity score model specification using Hosmer and Lemeshow’s goodness-of-fit test, a link test, and variance inflation factors (VIFs) to test for multicollinearity.

We implemented PSM using Stata 16 (43) and the kmatch command (44). We used kernel-based matching which includes all available observations and constructs a weighted average of counterfactuals for each observation in the treatment group. More information is taken from matches whose propensity scores are closer to each other and less information from matches whose propensity scores are further apart (41). We estimated the ATT using an Epanechnikov kernel and automatic bandwidth selection, meaning the bandwidth was determined by cross-validation using the propensity score (bandwidth=0.008) (44). We also imposed the common support condition to improve the quality of matches (45). Normal-based 95% confidence intervals and p values were estimated using bootstrapping with 500 replications.

#### Sensitivity analyses

We tested how robust our results were to different specifications of bandwidth values (0.01, 0.05) in the PSM models. We also tested PSM models without the common support condition. We then cross-validated our findings using an alternative approach; each wellbeing outcome was analyzed in a separate linear regression model, and all models are presented before and after adjustment for covariates.

Our definition of no arts group participation included those who reported participating both ‘never’ and ‘not in the last month’, meaning individuals who usually attend arts groups but had not been able to in the last month may have been wrongly categorized. We thus repeated our main analyses limiting the control group to those who never participated in arts groups, excluding individuals who reported not participating in the last month.

We also tested whether limiting the treatment group to individuals who participated in arts groups more frequently altered our findings. To do this, we repeated our main analyses with a new binary treatment indicator of no participation (never/not in the last month) versus participation weekly or more frequently. Individuals who participated monthly were excluded from this analysis.

Finally, we investigated whether associations between arts group participation and wellbeing were maintained longitudinally. In HRS, participants are eligible for the Psychosocial and Lifestyle Questionnaire every four years. Half of our sample were thus eligible to complete additional measures of wellbeing in 2018. We identified participants who completed all six measures of wellbeing in 2018 and repeated PSM models using arts group participation and covariates measured in 2014, with wellbeing four years later as the outcome.

## Results

In our sample (n=12,111), 10% reported participating in a community arts group in the last month. Before matching, there were differences across all covariates between those who did and did not participate in arts groups (Table 1). Reported life satisfaction (evaluative wellbeing), positive and negative affect (experienced wellbeing), and purpose in life, constraints and mastery (eudaimonic wellbeing) spanned the full range of potential scores before standardization (Table S1) and were moderately correlated (r=0.34-0.59; Figure S1).

**Table 1.** Demographic, socioeconomic, and health-related characteristics in the total sample and separately for those who did (treatment) and did not (control) participate in arts groups in the last month.

|  | Overall<br>(n=12,111) | Treatment<br>(n=1,252) | Control<br>(n=10,859) |
| --- | --- | --- | --- |
|  | N (%) |  |  |
| Gender |  |  |  |
| Women | 7168 (59%) | 820 (65%) | 6348 (58%) |
| Men | 4943 (41%) | 432 (35%) | 4511 (42%) |
| Race/ethnicity |  |  |  |
| White/Caucasian | 9048 (75%) | 805 (64%) | 8243 (76%) |
| Black/African American | 2093 (17%) | 366 (29%) | 1727 (16%) |
| Other (AI, AN, API, other) | 970 (8%) | 81 (6%) | 889 (8%) |
| Education |  |  |  |
| None | 1758 (15%) | 130 (10%) | 1628 (15%) |
| High school | 6376 (53%) | 564 (45%) | 5812 (54%) |
| College | 2639 (22%) | 341 (27%) | 2298 (21%) |
| Postgraduate | 1338 (11%) | 217 (17%) | 1121 (10%) |
| Marital status |  |  |  |
| Married | 7075 (58%) | 710 (57%) | 6365 (59%) |
| Divorced/Separated | 2137 (18%) | 213 (17%) | 1924 (18%) |
| Widowed | 2244 (19%) | 263 (21%) | 1981 (18%) |
| Never married | 655 (5%) | 66 (5%) | 589 (5%) |
| Employment status |  |  |  |
| Employed | 4055 (33%) | 462 (37%) | 3593 (33%) |
| Unemployed | 281 (2%) | 25 (2%) | 256 (2%) |
| Disabled | 1209 (10%) | 108 (9%) | 1101 (10%) |
| Retired | 5845 (48%) | 598 (48%) | 5247 (48%) |
| Homemaker | 721 (6%) | 59 (5%) | 662 (6%) |
| Neighborhood safety |  |  |  |
| Excellent | 3851 (32%) | 397 (32%) | 3454 (32%) |
| Very good | 4131 (34%) | 438 (35%) | 3693 (34%) |
| Good | 2709 (22%) | 269 (21%) | 2440 (22%) |
| Fair | 1159 (10%) | 119 (10%) | 1040 (10%) |
| Poor | 261 (2%) | 29 (2%) | 232 (2%) |
| Socializing frequency |  |  |  |
| Less than once a year | 363 (3%) | 17 (1%) | 346 (3%) |
| 1-2 per year | 498 (4%) | 27 (2%) | 471 (4%) |
| Every few months | 1031 (9%) | 67 (5%) | 964 (9%) |
| 1-2 per month | 2923 (24%) | 274 (22%) | 2649 (24%) |
| 1-2 per week | 4508 (37%) | 512 (41%) | 3996 (37%) |
| 3+ per week | 2788 (23%) | 355 (28%) | 2433 (22%) |
| Long term condition |  |  |  |
| None | 1372 (11%) | 173 (14%) | 1199 (11%) |
| One or more | 10739 (89%) | 1079 (86%) | 9660 (89%) |
|  | Mean (SD) |  |  |
| Age (years) | 68.09 (10.32) | 67.25 (9.92) | 68.19 (10.36) |
| Household income (USD) | 73,541 (10,5145) | 80,513 (11,4485) | 72,737 (10,3990) |
| Difficulties with ADLs | 0.30 (0.83) | 0.16 (0.58) | 0.31 (0.85) |
| Difficulties with IADLs | 0.24 (0.71) | 0.14 (0.56) | 0.25 (0.73) |
| Cognition | 9.75 (3.26) | 10.20 (3.32) | 9.70 (3.25) |

Note. AI: American Indian. AN: Alaska Native. API: Asian or Pacific Islander.

The propensity score model fitted the data well (χ^2^(53)=512.18, p<0.001, pseudo R^2^=0.06) with no evidence of misspecification or multicollinearity (VIF mean=1.30, range=1.02-1.75). Using this propensity score resulted in high quality matching (Figure S2) and corrected the balance of covariates between those who did and did not participate in arts groups (standardized mean difference range=0.0003-0.01; Table S2).

In the matched sample, we found evidence that arts group participation was associated with several aspects of wellbeing (Table 2). The strongest evidence was for the association with positive affect (experienced wellbeing), as arts group participation was associated with a 0.19 standard deviation higher positive affect compared to not participating in arts groups (ATT=0.19, 95% CI=0.13-0.24). Participating in arts groups was also associated with higher life satisfaction (evaluative wellbeing; ATT=0.10, 95% CI=0.05-0.16), purpose in life (eudaimonic wellbeing; ATT=0.08, 95% CI=0.02-0.14), and perceived mastery (eudaimonic wellbeing; ATT=0.08, 95% CI=0.02-0.13) than not participating. However, there was no evidence that arts group participation was associated with negative affect (experienced wellbeing) or perceived constraints on personal control (eudaimonic wellbeing).

**Table 2.**
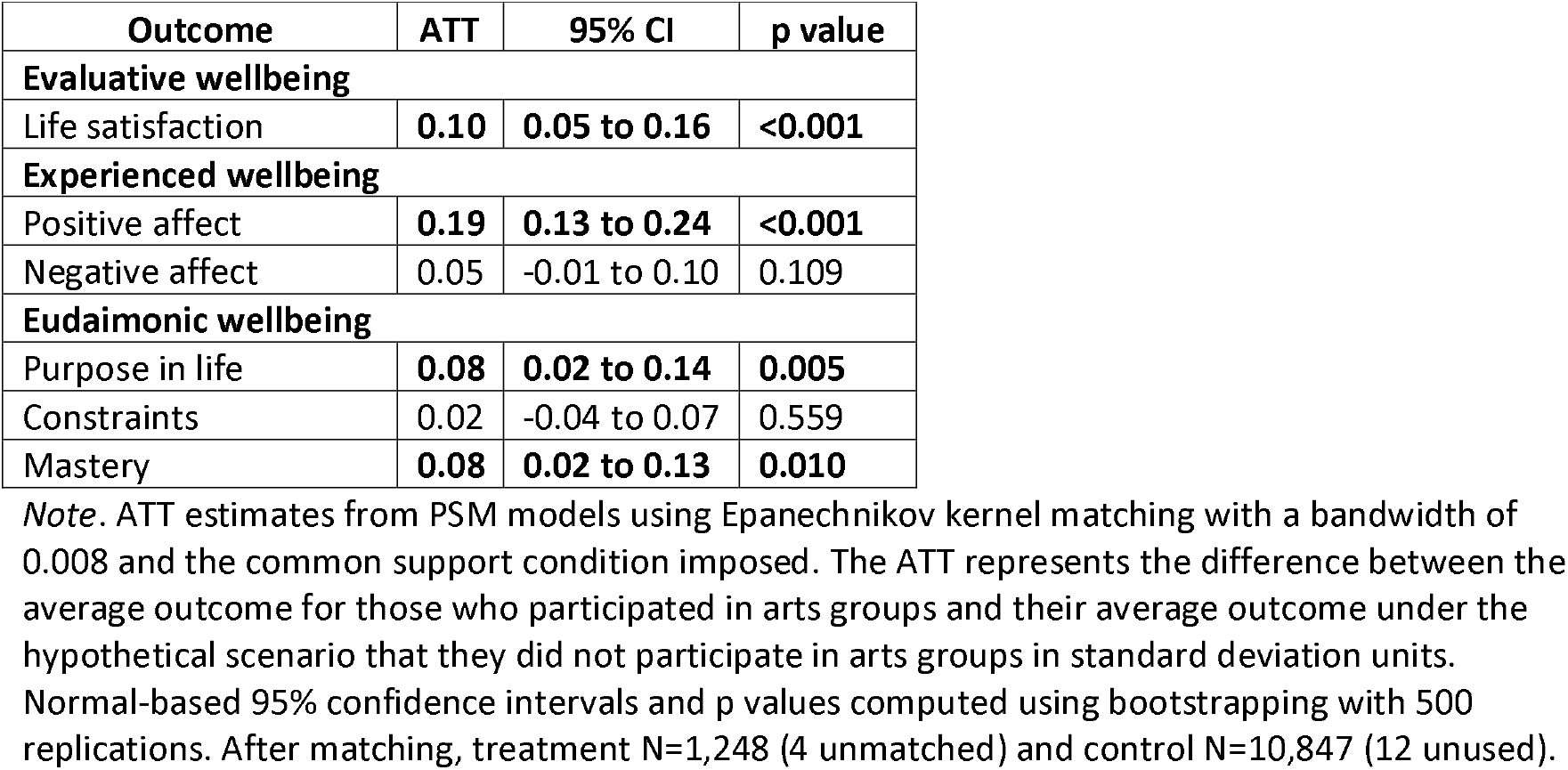
Associations between arts group participation (not in the last month vs once or more in the last month) and standardized wellbeing outcomes using propensity score matching.

| **Outcome** | **ATT** | **95% CI** | **p value** |
| --- | --- | --- | --- |
| **Evaluative wellbeing** |  |  |  |
| Life satisfaction | **0.10** | **0.05 to 0.16** | **<0.001** |
| **Experienced wellbeing** |  |  |  |
| Positive affect | **0.19** | **0.13 to 0.24** | **<0.001** |
| Negative affect | 0.05 | -0.01 to 0.10 | 0.109 |
| **Eudaimonic wellbeing** |  |  |  |
| Purpose in life | **0.08** | **0.02 to 0.14** | **0.005** |
| Constraints | 0.02 | -0.04 to 0.07 | 0.559 |
| Mastery | **0.08** | **0.02 to 0.13** | **0.010** |

*Note.* ATT estimates from PSM models using Epanechnikov kernel matching with a bandwidth of 0.008 and the common support condition imposed. The ATT represents the difference between the average outcome for those who participated in arts groups and their average outcome under the hypothetical scenario that they did not participate in arts groups in standard deviation units. Normal-based 95% confidence intervals and p values computed using bootstrapping with 500 replications. After matching, treatment N=1,248 (4 unmatched) and control N=10,847 (12 unused).

### Sensitivity analyses

Our results were not substantially altered in sensitivity analyses using a kernel bandwidth of 0.01 or 0.05 or when the common support requirement was removed (Table S3). We then cross-validated our findings using linear regression models. In the fully adjusted regression models, there was similar evidence that participation in arts groups was associated with higher life satisfaction, positive affect, purpose in life, and mastery but not associated with negative affect or constraints (Table S4).

Next, we limited the control group to individuals who never participated in arts groups, meaning those who reported that they had not participated in the last month were excluded from analyses. This did not alter our findings, except that participating in arts groups was then associated with higher negative affect (ATT=0.06, 95% CI=0.01-0.12: Table S5). However, the evidence for this association was weak and the ATT very similar to the ATT in our main analyses. We then limited the treatment group to those with at least weekly participation in arts groups, meaning the treatment and control groups were more distinct. In the matched sample, participating in arts group weekly or more often was associated with higher positive affect (ATT=0.21, 95% CI=0.14-0.29), life satisfaction (ATT=0.12, 95% CI=0.06-0.19), and purpose in life (ATT=0.11, 95% CI=0.04-0.18) compared to not participating (Table S6). As with at least monthly participation, there was no evidence for associations with negative affect or constraints. In contrast to monthly participation, there was only very weak evidence that weekly arts group participation was associated with higher mastery (ATT=0.07, 95% CI=-0.01-0.14). However, the ATT was the same as for monthly participation so this could be a result of the reduction in sample size in this sensitivity analysis.

Finally, we investigated whether associations between arts group participation and wellbeing were maintained longitudinally. In total, 3,888 participants completed the arts group participation measure in 2014 and wellbeing outcomes in 2018. In the matched sample, participating in arts groups monthly or more often was associated with higher positive affect (ATT=0.24, 95% CI=0.14-0.35), life satisfaction (ATT=0.20, 95% CI=0.10-0.29), purpose in life (ATT=0.13, 95% CI=0.02-0.24), and mastery (ATT=0.11, 95% CI=0.001-0.22) four years later compared to not participating (Table S7).

## Discussion

This study explored the associations between participation in a local community arts group (such as a choir, dance, photography, theatre, or music group) and a range of wellbeing outcomes. After matching on a range of demographic, socioeconomic, and health-related factors, we found evidence that arts group participation was associated with higher levels of the positive elements of evaluative, experienced, and eudaimonic wellbeing. Participation in an arts group was most strongly associated with positive affect, life satisfaction, purpose in life, and perceived mastery both cross-sectionally and in longitudinal sensitivity analyses. Comparing our findings for monthly and weekly arts group participation suggests that there could be a dose-response relationship, as the associations were slightly larger for weekly then monthly participation. We did not find evidence that participation was associated with negative aspects of experienced wellbeing (negative affect) and eudaimonic wellbeing (perceived constraints on personal control).

Our findings are consistent with previous evidence that arts engagement is associated with enhanced experienced, evaluative, and eudaimonic wellbeing in older adults (20–22). We have built on this research by using more sophisticated statistical techniques, demonstrating that although arts group participation is associated with broader aspects of social and cultural capital and socioeconomic status (which are themselves associated with wellbeing) (10,27,29), the relationship also exists independent of these factors in older adults. To our knowledge, no previous studies have used propensity score matching to investigate associations between participation in arts groups and wellbeing in older adults. Studies using similar methods in younger adults have found inconsistent evidence for the association between arts engagement and wellbeing, which is likely a result of the measures of arts engagement used, as well as differences in follow-up periods (23,31,32).

The relationship between arts group participation and wellbeing is likely to be bidirectional, as wellbeing also predicts subsequent arts engagement (12). As this study is observational and uses cross-sectional data, we have not attempted to show the direction of this association. We have focused instead on showing the independence of the association from a range of sociodemographic and health-related covariates. However, in a sensitivity analysis (albeit with a substantially reduced sample size), we found evidence that the associations between arts group participation and wellbeing were maintained over four years. This could be the result of a number of mechanistic pathways, including increased cognitive stimulation, physical activity, social interactions, emotional bonding, collaborative learning, pursuit of collective goals, and self-esteem as a result of arts participation (14–17).

In this study, participating in arts groups was associated only with the positive aspects of experienced and eudaimonic wellbeing. It is unclear why this occurred, particularly as participation was associated with evaluative wellbeing. Positive and negative affect are separate domains, and not just opposite states of experienced wellbeing. Positive affect includes feeling enthusiastic, alert, and pleasurable engagement in activities, whereas negative affect generally includes distress, anger, disgust, and fear (46). Arts group participation may thus increase positive affective experiences, without decreasing negative affect resulting from other sources. However, this could still lead to an enhanced ratio of positive to negative affect that would improve experienced wellbeing overall, particularly given that negative affect may decline in older age (47,48).

In terms of eudaimonic wellbeing, arts group participation was associated with purpose in life and perceived mastery but not perceived constraints on personal control. Alongside the health-promoting activities involved in participation, the act of participating in arts groups may directly provide a sense of purpose for individuals. Perceived mastery and constraints, in contrast, are two aspects of perceived control (individual’s beliefs that they are able to influence their circumstances) (29). Mastery relates to beliefs about abilities to achieve desired outcomes whereas constraints are beliefs about having obstacles that interfere with goal achievement. Beliefs about mastery may be more likely to impact an individual’s ability to attain desired outcomes, whereas constraints could be more likely to influence their ability to control life circumstances (49,50). Participating in arts groups may therefore increase older adults’ self-efficacy, promoting beliefs that they can achieve their goals, without altering their beliefs about external factors that may influence their lives. It is also possible that this distinction reflects the bidirectional nature of the association between arts group participation and wellbeing, as individuals who feel more able to achieve their goals may be more likely to join groups.

This study has a number of strengths. HRS is a large nationally representative cohort of older adults in the US. The rich data allowed us to match participants on a large set of covariates, which minimized the risk of bias caused by unobserved heterogeneity. We used validated measures of six aspects of wellbeing and a clearly defined measure of arts engagement. However, our study also has several limitations. Our main analyses were cross-sectional, so we cannot interpret the direction of the relationship between participation in arts groups and wellbeing or infer causality. This remains to be explored in future studies of older adults. PSM cannot control for any unobserved factors which may have influenced both arts group participation and wellbeing. But, given the wide range of covariates we included in our models, any remaining unobserved heterogeneity should be relatively small. Additionally, monthly arts group participation was not very common, meaning a relatively small proportion of the sample (10%) was included in the treatment group. Despite this, using kernel-based matching meant that nearly all participants were retained in the PSM models. Although we tested more frequent (weekly) arts group participation in sensitivity analyses with comparable results, this threshold resulted in an even smaller treatment group. Future studies could include larger samples or more prevalent forms of arts engagement to confirm our findings. Furthermore, we recognize that we performed PSM using an overly simple race/ethnicity variable (White, Black, Other), as defined in the HRS public data. This approach conflates experiences across diverse racial/ethnic groups, which might be particularly problematic as these groups may not have equal access to community arts groups and race/ethnicity may also be associated with wellbeing (10,51). As in previous research, the majority of participants in this study were White. In addition, the way in which race/ethnicity was reported, using only participants’ primary race/ethnicity, led to the erasure of multiracial persons in this study. Future research should thus use more diverse samples and collect more detailed data on race/ethnicity, while considering that racial and ethnic discrimination is a psychosocial stressor for historically racialized populations that adversely affects mental health (51), and may only be alleviated by the eradication of racism (52).

In this study, participation in local community arts groups (e.g., choirs, dance, photography, theatre, music groups) was associated with higher levels of life satisfaction (evaluative wellbeing), positive affect (experienced wellbeing), and purpose in life and perceived mastery (eudaimonic wellbeing). When considered alongside previous evidence for associations between arts engagement and subsequent wellbeing in older adults, our findings highlight the importance of future research that investigates ways to promote and facilitate participation in community arts groups. This could help to promote healthy aging, enabling a growing segment of the population to lead healthier and more satisfying lives. This is particularly important given that wellbeing is likely to not only be a product of arts engagement, but also to contribute to future health-related behaviors. Arts engagement may therefore be part of a beneficial cycle of enhanced wellbeing in older adults.

## Data Availability

In this study, we combined five Health and Retirement Study public datasets (RAND HRS: Longitudinal File 2018 (V1); Detailed Imputations File 2018 (V1); 2014-2018 Fat Files).

https://hrsdata.isr.umich.edu/data-products/rand

## Acknowledgements

We thank Shanae Burch, thought leader on work at the intersections of the arts, equity, and public health in the US, for her comments on this manuscript. We also gratefully acknowledge the contribution of the HRS study participants.

