## Supplementary Materials for "Associations between participation in community arts groups and aspects of wellbeing in older adults in the United States: A propensity score matching analysis"

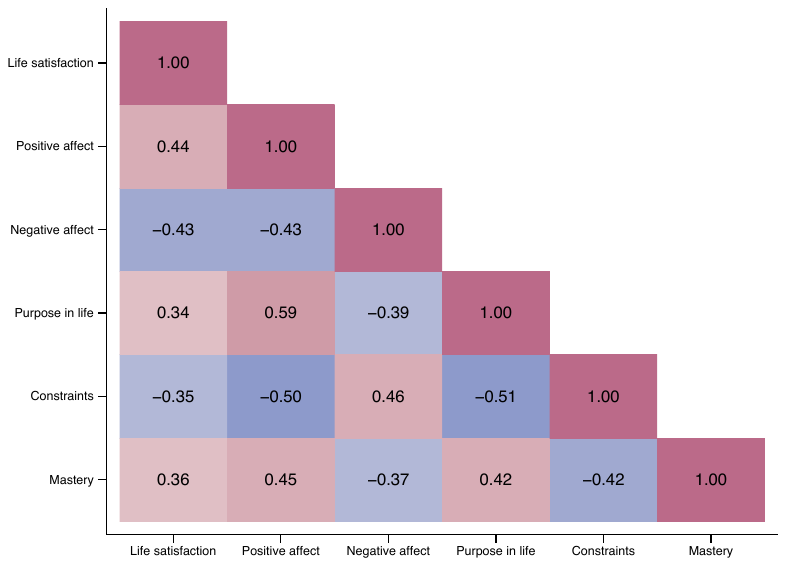

**Figure S1.** Heatmap showing the correlations between the five components of wellbeing.

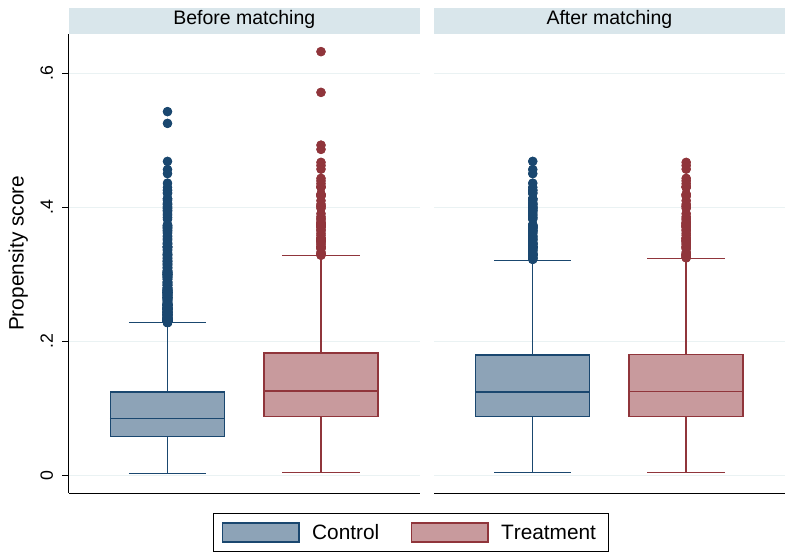

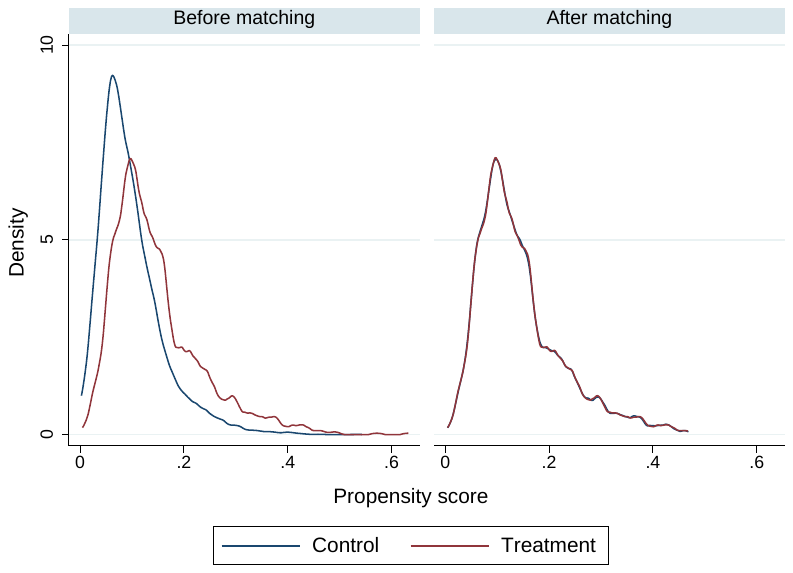

**Figure S2.** Propensity scores for participants who did not participate in arts groups (control) and those who participated in arts groups at least once in the last month (treatment), before and after propensity score matching.

**Table S1.** Mean wellbeing scores before standardization in the total sample and separately for those who did (treatment) and did not (control) participate in arts groups in the last month.

|  | **Overall**  (n=12,111) | | **Treatment**  (n=1,252) | | **Control**  (n=10,859) | |
| --- | --- | --- | --- | --- | --- | --- |
| **Outcome** | Range | Mean (SD) | Range | Mean (SD) | Range | Mean (SD) |
| **Evaluative wellbeing** | | | | | | |
| Life satisfaction | 1.00-7.00 | 5.00 (1.51) | 1.00-7.00 | 5.22 (1.42) | 1.00-7.00 | 4.98 (1.51) |
| **Experienced wellbeing** | | | | | | |
| Positive affect | 1.00-5.00 | 3.56 (0.81) | 1.00-5.00 | 3.81 (0.76) | 1.00-5.00 | 3.53 (0.81) |
| Negative affect | 1.00-5.00 | 1.75 (0.64) | 1.00-4.67 | 1.74 (0.62) | 1.00-5.00 | 1.76 (0.64) |
| **Eudaimonic wellbeing** | | | | | | |
| Purpose in life | 1.00-6.00 | 4.58 (0.95) | 2.14-6.00 | 4.81 (0.86) | 1.00-6.00 | 4.55 (0.95) |
| Constraints | 1.00-6.00 | 2.11 (1.15) | 1.00-6.00 | 1.99 (1.05) | 1.00-6.00 | 2.13 (1.16) |
| Mastery | 1.00-6.00 | 4.75 (1.13) | 1.00-6.00 | 4.91 (1.08) | 1.00-6.00 | 4.73 (1.13) |

**Table S2.** Balance of covariates between those who did (treatment) and did not participate in arts groups (control), and standardized differences between groups, before and after propensity score matching.

| **Variable** | **Raw** (N=12,111) | | | **Matched** (N=12,095) | | |
| --- | --- | --- | --- | --- | --- | --- |
|  | **Treatment** | **Control** | **Std diff** | **Treatment** | **Control** | **Std diff** |
|  | **Proportion** | | | | | |
| **Gender** | | | | | | |
| Men | 0.35 | 0.42 | -0.15 | 0.35 | 0.34 | 0.007 |
| **Race/ethnicity** | | | | | | |
| Black/African American | 0.29 | 0.16 | 0.32 | 0.29 | 0.29 | 0.007 |
| Other (AI, AN, API, other) | 0.06 | 0.08 | -0.07 | 0.06 | 0.06 | 0.003 |
| **Education** | | | | | | |
| High school | 0.45 | 0.54 | -0.17 | 0.45 | 0.45 | 0.01 |
| College | 0.27 | 0.21 | 0.14 | 0.27 | 0.27 | -0.002 |
| Postgraduate | 0.17 | 0.10 | 0.20 | 0.17 | 0.18 | -0.01 |
| **Marital status** | | | | | | |
| Divorced/Separated | 0.17 | 0.18 | -0.02 | 0.17 | 0.17 | 0.004 |
| Widowed | 0.21 | 0.18 | 0.07 | 0.21 | 0.21 | 0.0003 |
| Never married | 0.05 | 0.05 | -0.01 | 0.05 | 0.05 | 0.001 |
| **Employment status** | | | | | | |
| Unemployed | 0.02 | 0.02 | -0.02 | 0.02 | 0.02 | -0.004 |
| Disabled | 0.09 | 0.10 | -0.05 | 0.09 | 0.08 | 0.005 |
| Retired | 0.48 | 0.48 | -0.01 | 0.48 | 0.48 | -0.003 |
| Homemaker | 0.05 | 0.06 | -0.06 | 0.05 | 0.05 | 0.002 |
| **Neighborhood safety** | | | | | | |
| Very good | 0.35 | 0.34 | 0.02 | 0.35 | 0.35 | 0.006 |
| Good | 0.21 | 0.22 | -0.02 | 0.21 | 0.21 | 0.0006 |
| Fair | 0.10 | 0.10 | 0.00 | 0.09 | 0.10 | -0.006 |
| Poor | 0.02 | 0.02 | 0.01 | 0.02 | 0.02 | 0.005 |
| **Socializing frequency** | | | | | | |
| 1-2 per year | 0.02 | 0.04 | -0.12 | 0.02 | 0.02 | -0.002 |
| Every few months | 0.05 | 0.09 | -0.14 | 0.05 | 0.05 | -0.003 |
| 1-2 per month | 0.22 | 0.24 | -0.06 | 0.22 | 0.22 | 0.007 |
| 1-2 per week | 0.41 | 0.37 | 0.08 | 0.41 | 0.41 | -0.002 |
| 3+ per week | 0.28 | 0.22 | 0.14 | 0.28 | 0.28 | -0.0009 |
| **Long term condition** | | | | | | |
| One or more | 0.86 | 0.89 | -0.08 | 0.86 | 0.86 | 0.002 |
|  | **Mean** | | | | | |
| Age (years) | 67.25 | 68.19 | -0.09 | 67.25 | 67.34 | -0.01 |
| Household income (USD) | 80513.07 | 72737.16 | 0.07 | 80539.32 | 79869.10 | 0.006 |
| Difficulties with ADLs | 0.16 | 0.31 | -0.22 | 0.16 | 0.16 | -0.004 |
| Difficulties with IADLs | 0.14 | 0.25 | -0.16 | 0.14 | 0.14 | -0.003 |
| Cognition | 10.20 | 9.70 | 0.15 | 10.21 | 10.22 | -0.004 |

*Note.* Std diff: Standardized difference. AI: American Indian. AN: Alaska Native. API: Asian or Pacific Islander. Before matching, treatment N=1,252 and control N=10,859. After matching, treatment N=1,248 (4 unmatched) and control N=10,847 (12 unused).

**Table S3.** Alternative model specifications (kernel bandwidth of 0.01 or 0.05 or no common support requirement) for testing associations between participation in arts groups and standardized wellbeing outcomes using propensity score matching.

| **Outcome** | **Bandwidth = 0.01** | | | **Bandwidth = 0.05** | | | **Automatic bandwidth,**  **no common support** | | |
| --- | --- | --- | --- | --- | --- | --- | --- | --- | --- |
|  | ATT | 95% CI | p value | ATT | 95% CI | p value | ATT | 95% CI | p value |
| **Evaluative wellbeing** | | | | | | | | | |
| Life satisfaction | **0.10** | **0.05 to 0.16** | **<0.001** | **0.11** | **0.05 to 0.16** | **<0.001** | **0.10** | **0.05 to 0.16** | **<0.001** |
| **Experienced wellbeing** | | | | | | | | | |
| Positive affect | **0.19** | **0.13 to 0.24** | **<0.001** | **0.20** | **0.15 to 0.26** | **<0.001** | **0.19** | **0.13 to 0.24** | **<0.001** |
| Negative affect | 0.05 | -0.01 to 0.10 | 0.106 | 0.04 | -0.01 to 0.10 | 0.142 | 0.05 | -0.01 to 0.10 | 0.109 |
| **Eudaimonic wellbeing** | | | | | | | | | |
| Purpose in life | **0.08** | **0.02 to 0.14** | **0.005** | **0.10** | **0.04 to 0.16** | **0.001** | **0.08** | **0.02 to 0.14** | **0.005** |
| Constraints | 0.02 | -0.04 to 0.07 | 0.561 | 0.003 | -0.05 to 0.06 | 0.918 | 0.02 | -0.04 to 0.07 | 0.561 |
| Mastery | **0.08** | **0.02 to 0.13** | **0.009** | **0.09** | **0.03 to 0.14** | **0.003** | **0.08** | **0.02 to 0.13** | **0.009** |

*Note*. For all models, normal-based 95% confidence intervals and p values computed using bootstrapping with 500 replications. For bandwidth of 0.01, treatment N=1,248 (4 unmatched) and control N=10,847 (12 unused). For bandwidth of 0.05, treatment N=1,250 (2 unmatched) and control N=10,849 (10 unused). For automatic bandwidth (0.008) and no common support, treatment N=1,248 (4 unmatched) and control N=10,857 (2 unused).

**Table S4.** Linear regression models testing associations between participation in arts groups and standardized wellbeing outcomes.

| **Outcomes** | **Unadjusted** | | | **Adjusted** | | |
| --- | --- | --- | --- | --- | --- | --- |
|  | Coef | 95% CI | p value | Coef | 95% CI | p value |
| **Evaluative wellbeing** | | | | | | |
| Life satisfaction | **0.16** | **0.1 to 0.22** | **<0.001** | **0.10** | **0.05 to 0.16** | **<0.001** |
| **Experienced wellbeing** | | | | | | |
| Positive affect | **0.35** | **0.29 to 0.4** | **<0.001** | **0.19** | **0.14 to 0.25** | **<0.001** |
| Negative affect | -0.02 | -0.08 to 0.04 | 0.561 | 0.04 | -0.01 to 0.1 | 0.125 |
| **Eudaimonic wellbeing** | | | | | | |
| Purpose in life | **0.26** | **0.21 to 0.32** | **<0.001** | **0.09** | **0.04 to 0.15** | **0.001** |
| Constraints | **-0.12** | **-0.18 to -0.06** | **<0.001** | 0.01 | -0.04 to 0.07 | 0.615 |
| Mastery | **0.16** | **0.1 to 0.22** | **<0.001** | **0.08** | **0.02 to 0.14** | **0.007** |

*Note.* Each outcome was tested in a separate linear regression model. Models were adjusted for all covariates: age, gender, ethnicity, education, marital status, employment status, household income, neighborhood safety, socializing frequency, long term conditions, difficulties with ADLs and IADLS and cognition (see main text for further details).

**Table S5.** Associations between monthly participation in arts groups (never vs at least once in the last month) and standardized wellbeing outcomes using propensity score matching.

| **Outcome** | **ATT** | **95% CI** | **p value** |
| --- | --- | --- | --- |
| **Evaluative wellbeing** | | | |
| Life satisfaction | **0.11** | **0.06 to 0.17** | **<0.001** |
| **Experienced wellbeing** | | | |
| Positive affect | **0.21** | **0.16 to 0.27** | **<0.001** |
| Negative affect | **0.06** | **0.01 to 0.12** | **0.032** |
| **Eudaimonic wellbeing** | | | |
| Purpose in life | **0.10** | **0.05 to 0.16** | **<0.001** |
| Constraints | 0.01 | -0.04 to 0.07 | 0.632 |
| Mastery | **0.09** | **0.04 to 0.15** | **0.002** |

*Note*. ATT estimates from PSM models using Epanechnikov kernel matching with a bandwidth of 0.01 and the common support condition imposed. Normal-based 95% confidence intervals and p values computed using bootstrapping with 500 replications. After matching, treatment N=1,250 (2 unmatched) and control N=9,082 (18 unused).

**Table S6.** Associations between weekly participation in arts groups (none vs weekly or more often) and standardized wellbeing outcomes using propensity score matching.

| **Outcome** | **ATT** | **95% CI** | **p value** |
| --- | --- | --- | --- |
| **Evaluative wellbeing** | | | |
| Life satisfaction | **0.12** | **0.06 to 0.19** | **<0.001** |
| **Experienced wellbeing** | | | |
| Positive affect | **0.21** | **0.14 to 0.29** | **<0.001** |
| Negative affect | 0.01 | -0.06 to 0.08 | 0.796 |
| **Eudaimonic wellbeing** | | | |
| Purpose in life | **0.11** | **0.04 to 0.18** | **0.002** |
| Constraints | -0.01 | -0.08 to 0.05 | 0.694 |
| Mastery | 0.07 | -0.01 to 0.14 | 0.080 |

*Note*. ATT estimates from PSM models using Epanechnikov kernel matching with a bandwidth of 0.006 and the common support condition imposed. Normal-based 95% confidence intervals and p values computed using bootstrapping with 500 replications. After matching, treatment N=703 (2 unmatched) and control N=10,668 (191 unused).

**Table S7.** Longitudinal associations between monthly participation in arts groups and standardized wellbeing outcomes, measured four years later, using propensity score matching.

| **Outcome** | **ATT** | **95% CI** | **p value** |
| --- | --- | --- | --- |
| **Evaluative wellbeing** | | | |
| Life satisfaction | **0.20** | **0.10 to 0.29** | **<0.001** |
| **Experienced wellbeing** | | | |
| Positive affect | **0.24** | **0.14 to 0.35** | **<0.001** |
| Negative affect | 0.03 | -0.07 to 0.14 | 0.566 |
| **Eudaimonic wellbeing** | | | |
| Purpose in life | **0.13** | **0.02 to 0.24** | **0.016** |
| Constraints | -0.04 | -0.14 to 0.07 | 0.493 |
| Mastery | **0.11** | **0.001 to 0.22** | **0.048** |

*Note*. ATT estimates from PSM models using Epanechnikov kernel matching with a bandwidth of 0.02 and the common support condition imposed. Normal-based 95% confidence intervals and p values computed using bootstrapping with 500 replications. After matching, treatment N=425 (0 unmatched) and control N=3,360 (103 unused).
